# Breastfeeding support for mothers of low birth weight infants using mother-to-mother peers in rural western Kenya - a feasibility study

**DOI:** 10.1101/2023.01.23.23284905

**Authors:** Fiona M Dickinson, Florence Achieng, Alloys K’Oloo, Iwaret Otiti, Linda Tindi, Mwanamvua Boga, Mary Kimani, Laura Kiige, Kathy Mellor, Stephanie Dellicour, Hellen C. Barsosio, Simon Kariuki, Helen M Nabwera

## Abstract

The majority of the 2.4 million neonates (infants<28 days) who died in 2020 were born weighing <2500g i.e. low birth weight (LBW). In Africa, approximately 1 in 10 neonates are LBW. The majority of those who survive beyond the neonatal period are undernourished, have neuro-developmental impairment, or die before their second birthday. Unaddressed feeding difficulties contribute significantly to these adverse outcomes. This study assessed the feasibility and acceptability of using trained mother-to-mother peers (peer-mothers) to deliver breastfeeding support to mothers of LBW infants in healthcare facilities in rural Kenya.

A mixed methods approach was employed, using structured observations, and pre- and post-intervention semi-structured interviews, with mothers, healthcare providers and peer-mothers. Six trained peer-mothers delivered the interventions to the mother-LBW infant pairs across eight healthcare facilities in Homa Bay County. Descriptive statistics were used to analyse the quantitative data and thematic analysis for the qualitative data.

From September-November 2021, 23 mothers and 26 LBW infants were recruited and received the intervention. All infants were born in a hospital, the median age of the infants was 1 day (Interquartile range, IQR 1,3), birth weight 2100g (IQR 1900, 2260) and recorded gestation 34 weeks (IQR 34,36). Although all mothers looked well, 4 (17%) showed no signs of bonding with their infant. One infant was too weak to suckle and was referred to healthcare providers. Key themes were challenges with infant feeding decision-making among mothers of LBW infants, community misconceptions of recommended infant feeding practices for LBW infants, and the integral role of peer-mothers in maternity units.

Facility-based, breastfeeding peer support for LBW infants was feasible and acceptable in the context of resource constraints. It could improve uptake of appropriate infant feeding practices among these vulnerable infants and enhance their post-discharge survival and growth outcomes. This strategy warrants further evaluation in a larger study.

## Introduction

In 2020, 2.4 million neonates (infants <28 days) died worldwide, accounting for nearly half of under-5 deaths [1]. Despite significant global efforts and gains in addressing under-5 mortality, neonatal mortality has declined slowly, particularly in sub-Saharan Africa. The Sustainable Development Goal (SDG) target of reducing neonatal mortality rate to < 12 per 1,000 live births by 2030, is unlikely to be achieved by many sub-Saharan African countries [2]. Transformative and sustainable strategies are therefore required to address neonatal health problems.

Low birth weight (LBW, <2500g) infants include those who are preterm (<37 weeks gestation) and/or intrauterine growth-restricted infants [3]. They have the highest risk of mortality both in the neonatal and post-neonatal period [4,5]. In addition, survivors often have long-standing unaddressed nutritional problems [6,7,8]. Globally, approx. 20 million LBW infants are born annually, and over 90% are born in low-and middle-income countries (LMICs), such as Kenya. A key challenge in these countries is the inadequate prioritisation of maternal and newborn health and, therefore, resource allocation, compounded by the limited progress in empowering women [9,10,11].

WHO/UNICEF recommend that mothers are supported to initiate breastfeeding within an hour of delivery and avoid any prelacteal feeds that are harmful and often preclude newborns from establishing breastfeeding [12]. Thereafter, infants should be exclusively breastfed for six months [12]. These are key components of the WHO/UNICEF Baby Friendly Hospital Initiative that many hospitals in Kenya have signed up for but cannot sustain due to resource constraints [13]. Unfortunately, optimal breastfeeding is rarely achieved among LBW infants in LMICs, largely due to the limited support that is available for mothers in the immediate (<24 hours) and early (from 2-7 days) postnatal period [14,15]. In Kenya, postnatal care focuses on the management of acute and life-threatening problems with insufficient time and resources allocated to providing support for breastfeeding and infant care practices to caregivers - a vital component of addressing preventable deaths among the most vulnerable newborns.

Mother-to-mother peer support for LBW infants (peer-mothers) is a strategy that we have co-developed and explored in a community setting, where mothers who have experience in caring for a LBW infant are trained and mentored to provide breastfeeding and other key infant care practices in the postnatal period [16]. In the context of under-resourced maternal and newborn healthcare services, peer-mothers could play a critical role in supporting mothers of LBW infants to optimise their breastfeeding practices prior to discharge, with implications for their post-discharge outcomes [17]. The aim of this study was, therefore, to explore the feasibility and acceptability of using trained peer-mothers to deliver breastfeeding support to mothers of LBW infants in healthcare facilities in rural, western Kenya.

## Methods

### Setting

The study was conducted in eight healthcare facilities in Homa Bay County, western Kenya. Nearly two-thirds of the population of Homa Bay County live in extreme poverty, i.e. <1 USD per day [18]. It also has the highest rate of HIV in Kenya, with an estimated 25% prevalence among women aged 15-49 years [19]. Although the neonatal mortality rate is yet to be calculated for this County, the national estimate is 20 per 1,000 live births [20], which is higher than the global estimate of 17 per 1,000 live births [1]. Approximately 6% of infants are LBW with high rates of undernutrition in under 5’s: approximately 15% underweight, 25% stunted and 4% wasted [21]. Only 35% of infants under six months are exclusively breastfed [21].

### Study design

This was a quasi-experimental study that employed both qualitative and quantitative data collection methods [22]. This mixed methods approach enabled us to explore in-depth the factors that influence the feasibility and acceptability of using peer-mothers to support mothers of LBW infants to establish optimal breastfeeding practices, in healthcare facilities in Homa Bay County, western Kenya [22]. The study intervention comprised the trained peer-mothers providing breastfeeding support to mothers of LBW infants in the postnatal wards and clinics across eight healthcare facilities in Homa Bay County. Debriefing sessions with the peer-mothers, by the healthcare providers and the study team, took place every two weeks or more regularly as required.

### Participants and sampling

The mothers of LBW infants were recruited from the postnatal wards and clinics of the eight healthcare facilities in Homa Bay County, that were purposively selected in view of their varied resources and patient populations (county & sub-county level and mission hospitals as well as health centres). Inclusion criteria for the mother-LBW infant pairs were that the LBW infant was alive at birth, were < 28 days old, and the mother was ≥15 years old.

Recruitment of the peer-mothers was conducted using a 2-step process. Initially, 17 peer-mothers, who had experience of caring for a LBW infant, had completed primary school education and were recommended by community health workers, were invited to take part in a four-day breastfeeding support training package. The training package was co-developed by child health and lactation experts from Kenya and the United Kingdom [16], based on international recommendations [23,24]. The key components of the 4-day training package are shown in **Figure 1** and included communication skills, breastfeeding support, Kangaroo Mother Care (KMC), hygiene, and identification of danger signs.

**Figure 1:**
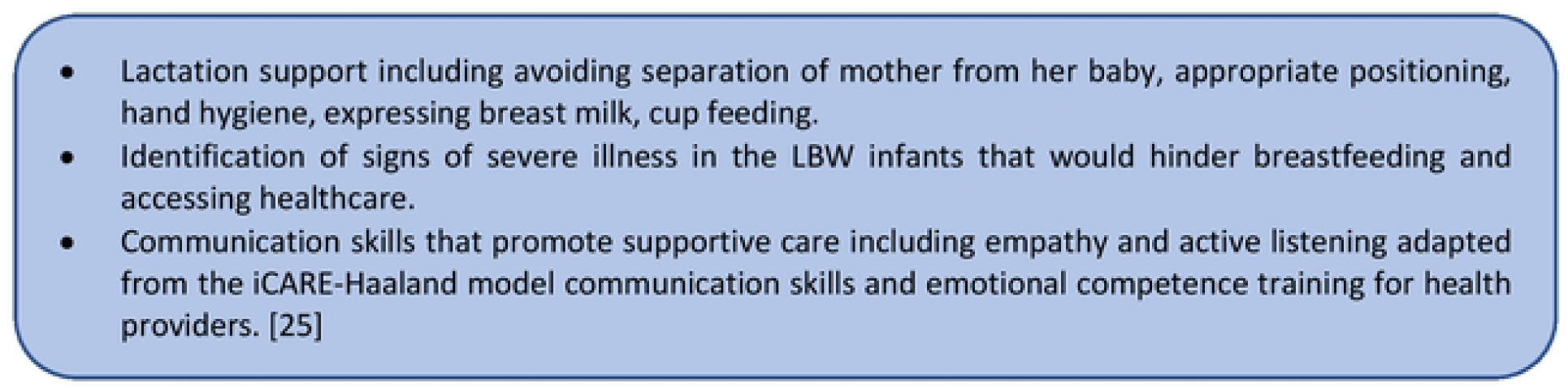
Key components of the training package for peer-mothers

At the end of the training, competency-based scenarios were used to select 10 (out of 17, 59%) peer-mothers who achieved the minimum standards of knowledge and skills required to deliver the intervention. However, due to delays caused by the health workers’ strikes, and in the absence of additional project funding, four peer-mothers were transferred to provide support with follow-up for an infant nutrition supplementation trial [26]. Therefore, six peer-mothers delivered breastfeeding support to the mother-LBW infant pairs, working alongside healthcare providers.

Purposive sampling was used to identify mothers who could potentially benefit from the intervention as they had given birth to a LBW infant (baseline) and those who had received the intervention and appeared to be progressing well with breastfeeding their LBW infant vs those whose LBW infant was not thriving (post-intervention) [27]. In addition, purposive sampling was used to identify healthcare providers working on the postnatal wards or newborn units with characteristics of interest including length of service (<5 years of service vs ≥ 5 years of service in the healthcare facility), having been trained on breastfeeding support strategies for LBW (yes vs no), and involvement in a leadership role within the healthcare facility (leadership vs non-leadership); both at baseline and post-intervention. The aim was to interview all peer-mothers who had delivered the intervention (5, 83% were available for the interview at the end of the study).

### Sample size

#### Quantitative

Based on the 2019 hospital statistics from Homa Bay County Referral Hospital, we estimated that approximately eight LBW infants per month would be born alive across the healthcare facilities. Over the three months of data collection, we therefore, expected a sample size of at least 24 mother-LBW infant pairs to describe breastfeeding counselling availability, timing and utility at baseline and after implementing peer-mother support in the healthcare facilities. As this study was not testing a hypothesis, no sample size calculation was done.

#### Qualitative

Twelve semi-structured interviews were conducted with mothers of LBW infants and healthcare providers at baseline (pre-intervention) to explore their perspectives and experiences of breastfeeding support, and 12 post-intervention interviews with mothers of live LBW newborns, healthcare providers and peer-mothers to explore their perspectives of the breastfeeding support intervention. These enabled us to achieve “data saturation” where no more new information emerged from additional interviews [28].

Twenty-six interviews were conducted, 10 pre-intervention and 16 post-intervention. These comprised 11 with mothers of LBW infants, 10 with healthcare providers, and 5 with peer-mothers (**Table 1**).

**Table 1.** Participants for the semi-structured interviews

|  | Pre-intervention<br>interviews | Post-intervention<br>interviews |
| --- | --- | --- |
| **Mothers of LBW infants** | 4 | 7 |
| **Peer-mothers** | - | 5 |
| **Healthcare providers** | 6 | 4 |

### Data collection

#### Quantitative

Quantitative data were collected on peer-mother activities (i.e. number of mother-infant pairs they supported over the study period and retention of mothers of LBW infants). Structured observations were also carried out by research assistants, to describe how the peer-mothers assessed the infants, with the key areas being the mother’s interactions with their LBW infant, and their infant feeding practices.

#### Qualitative

At baseline semi-structured interviews with mothers of LBW infants and healthcare providers were conducted to explore their experiences of receiving or perspectives of breastfeeding support in the healthcare facilities. This helped to refine the peer-mother breastfeeding training package by providing more context-relevant details that were not considered in the original version that focused on community-based delivery of the intervention.

Post-intervention, semi-structured interviews were conducted in the 0-7 days after birth, with mothers of LBW infants who received peer-mother support, as well as with healthcare providers and peer-mothers, to explore their experiences and acceptability of the intervention.

### Data Management and Analysis

#### Quantitative

Data were collected electronically using password-protected tablets and the Survey CTO software (https://www.surveycto.com). Anonymised data were transmitted via a secure connection to the Liverpool School of Tropical Medicine (LSTM) server. Data were then imported from excel to Stata 15 (StataCorp., Texas, USA) for analysis using descriptive statistics. This included summary statistics – mean (standard deviation), median (interquartile range), number (percentage), to describe the recruitment, training, characteristics, activity and retention of peer-mothers and the baseline characteristics of the mother-LBW infant pairs. The following indicators were explored to describe the feasibility of delivering this breastfeeding support using peer-mothers: number of peer-mothers identified who were recruited and completed the training; number of peer-mothers who engaged with debriefing sessions; and number of peer-mothers who continued to deliver the intervention throughout the study period.

#### Qualitative

Semi-structured interviews were recorded and stored on passcode-locked digital voice recorders. The interviews were transcribed and translated into English by one of the research assistants, a native speaker of both Kiswahili and Dholuo, the language in which the interviews were conducted. The study used a thematic analysis method [29] to explore data from the interviews and draw both descriptive and explanatory conclusions about their experience of breastfeeding support pre- and post-peer-mother intervention. The study team familiarised themselves with the data by re-reading the transcripts to identify recurring issues, inconsistencies, and possible categories, creating a coding framework and then coding using a mixture of deductive codes from our topic guide and inductive codes emerging from the data, which we then applied to all transcripts. The codes were then searched to identify themes, which were reviewed and defined. The coding process was an iterative process involving changing and adding to them as more data became available. The final coding framework developed was used to code the entire data set using the NVivo 11 software (QSR International Pty Ltd). Throughout this process, the study ensured peer review of transcripts and codes to increase reliability and reduce inconsistencies.

### Ethical statement

Ethics approval for the study was obtained from the Kenya Medical Research Institute’s Scientific and Ethics Review Unit (Nairobi, Kenya) (ref: KEMRI/SERU/CGHR/05-03-371/4200) and the LSTM Research Ethics Committee (UK) (ref: 21/026). Written, informed consent was obtained from all participants prior to data collection, including mothers of low birth weight infants. Due to high levels of illiteracy in the study communities, participants incapable of providing written informed consent were verbally guided through the study consent procedures and indicated their consent with a thumbprint. The study was conducted during the COVID-19 pandemic and study activities were done in accordance with the Kenya, Ministry of Health COVID-19 guidelines.

## Results

### Demographics

From September to November 2021, 23 mother-LBW infant pairs, including 3 sets of twins (26 infants in total), were recruited and received the intervention. The median maternal age was 25 years (interquartile range, IQR 22, 29), and all infants were delivered in a hospital. The median age of the infants was 1 day (IQR 1,3), median birth weight was 2100g (IQR 1900, 2260), and median gestation was 34 weeks (IQR 34,36).

### Quantitative findings

The structured observations covered the peer-mothers monitoring 5 key areas: maternal and infant well-being, infant positioning, attachment and suckling during breastfeeding.

All peer-mothers observed the mothers and their LBW infants’ well-being (n=23). Although most mothers looked relaxed and calm (n=19, 83%), 4 (17%) showed no signs of bonding with their infants, including lack of interaction with their infant or attention paid to breastfeeding cues **(Figure 2)**.

**Figure 2:**
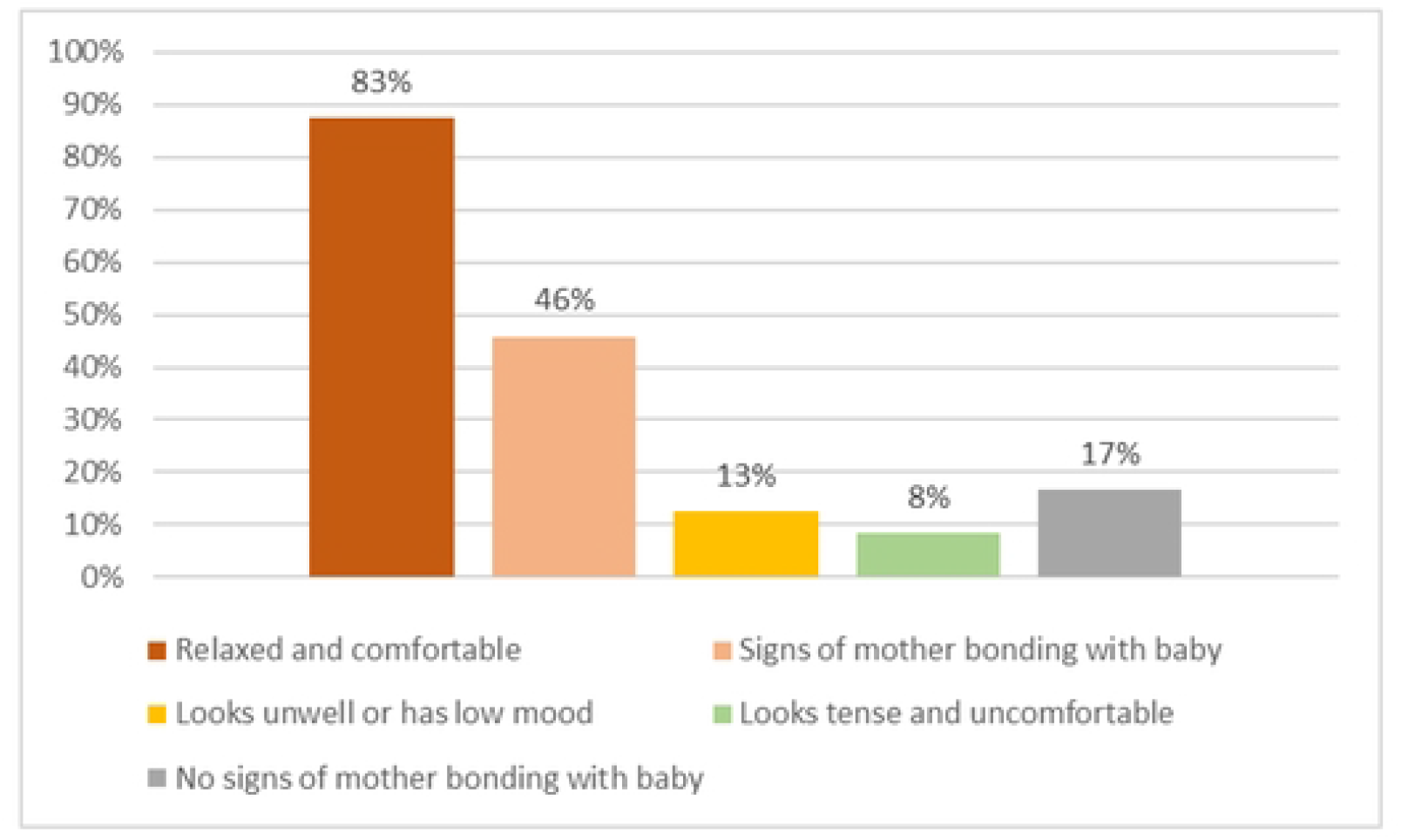
Exploration of maternal well-being

The majority of infants were alert and able to breastfeed (19, 83%) and were rooting for the breast (n=22, 96%), although one infant (4%) was noted to be restless **(Figure 3)**.

**Figure 3:**
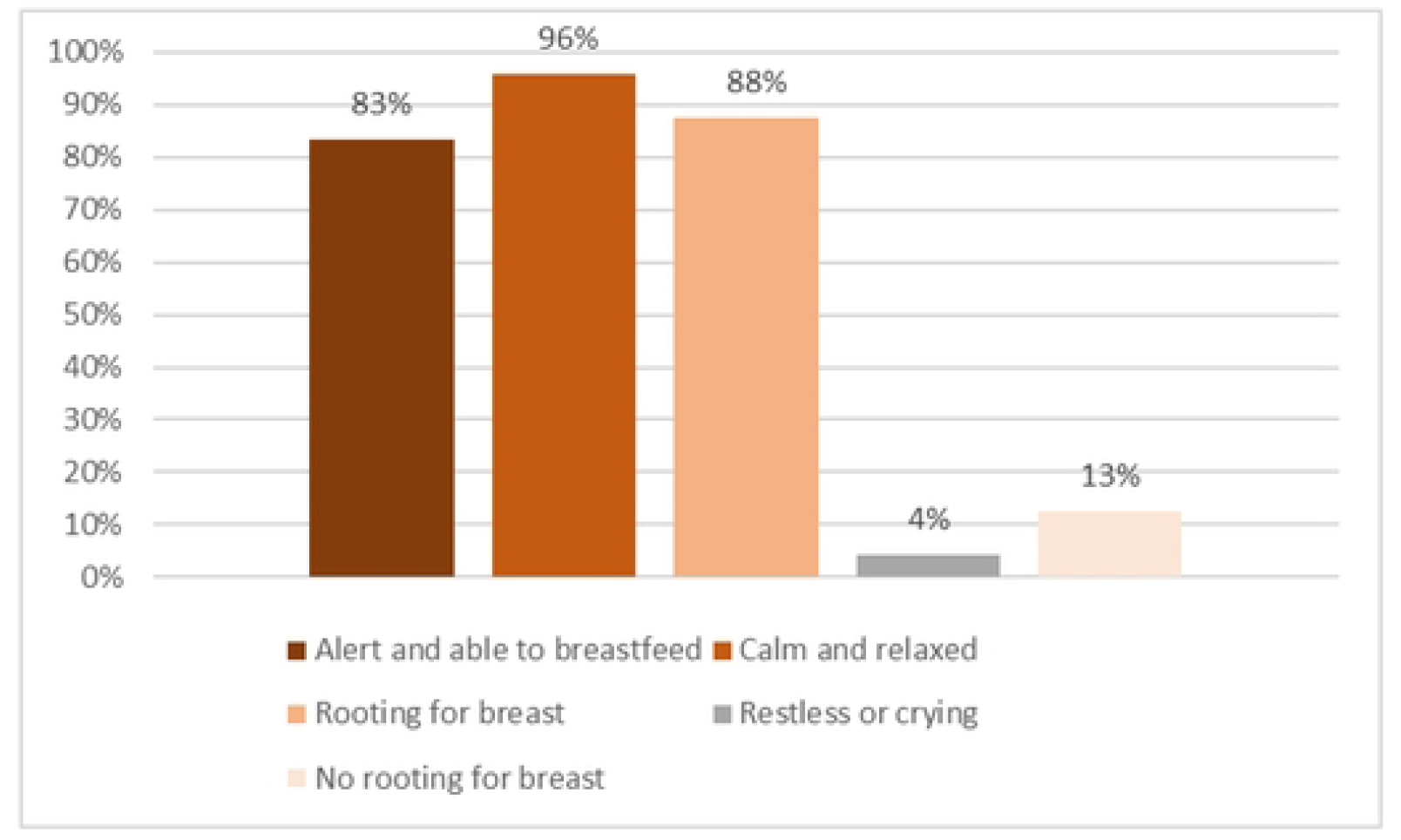
Exploration of low-birth-weight infants’ well-being

Of the 23 mother-infant pairs, most infants were held appropriately by their mothers (n=21, 91%) but less than half positioned their infants appropriately at the breast i.e. “approach breast nose to nipple” (10, 43%) **(Figure 4)**.

**Figure 4:**
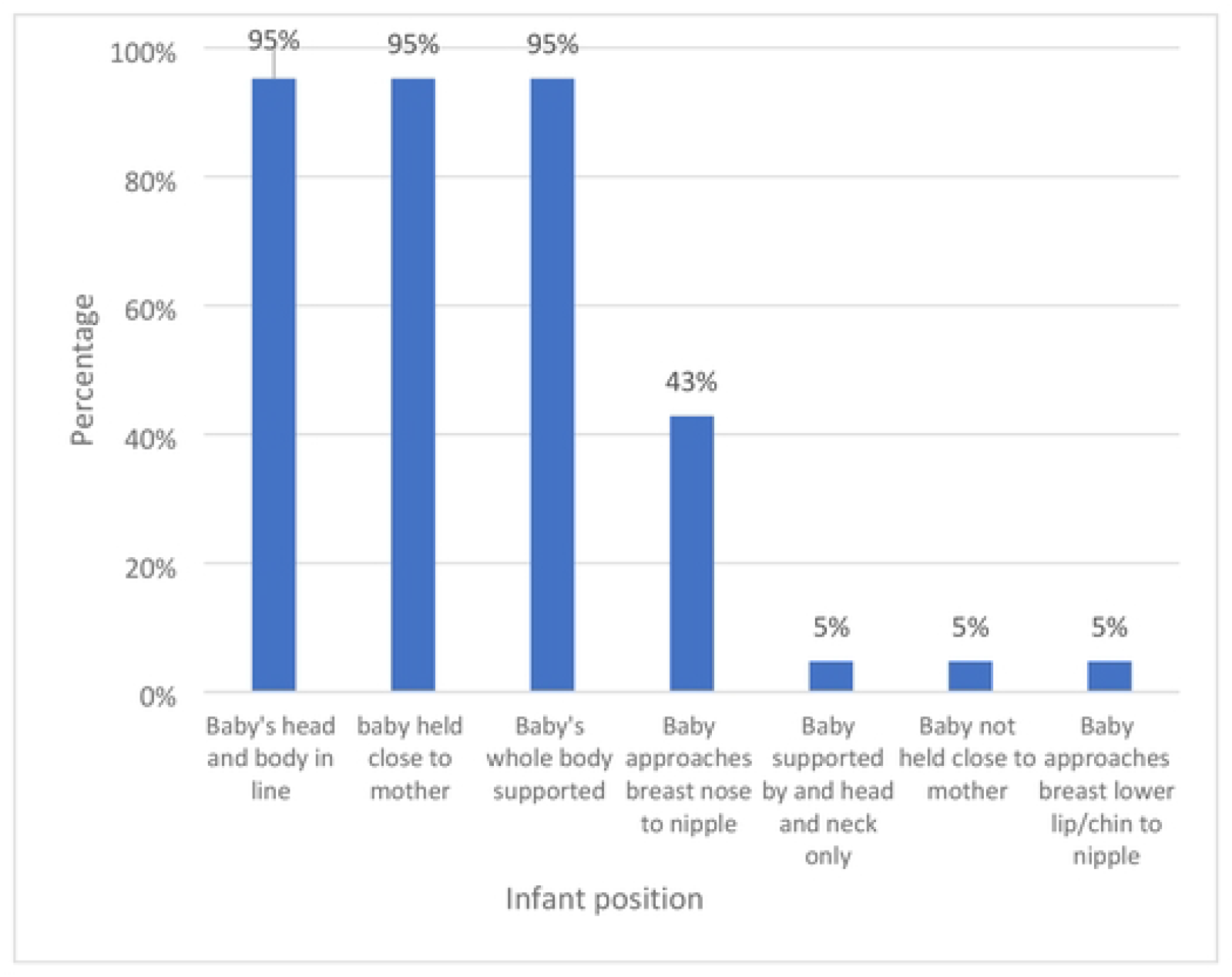
Position of the infant during breastfeeding

As part of the support intervention, the peer-mothers emphasised skin-to-skin contact between the LBW-mothers and their infants during breastfeeding. Of the 15 infants observed attaching to mothers’ breasts, more than a quarter (n=6, 27%) had inadequate attachment during the feed **(Figure 5)**.

**Figure 5:**
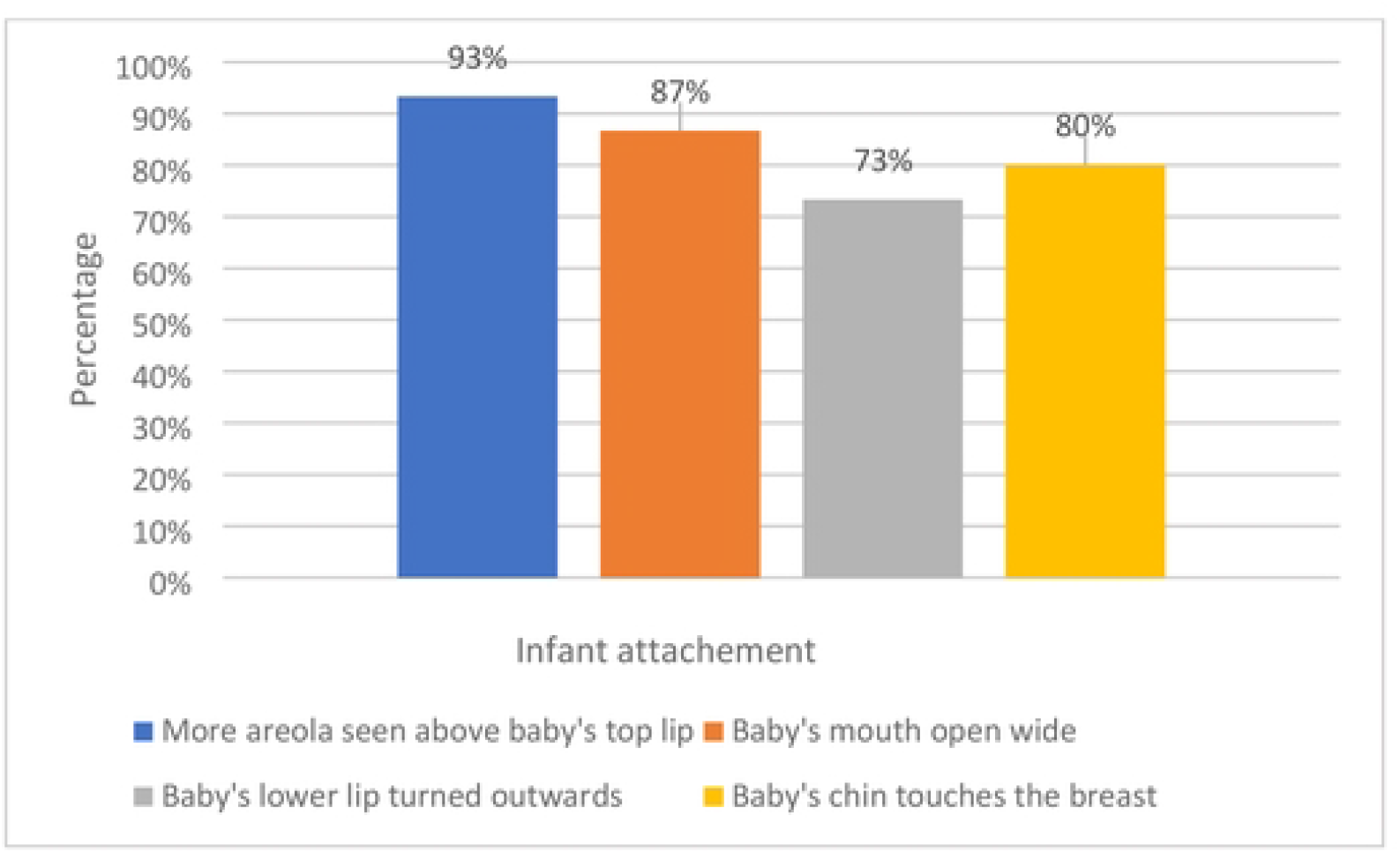
Infant attachment at mother’s breast during feeding

Infants were observed suckling among only 13(57%) of mother-infant pairs. Of these, all of them had slow deep sucks at one point, but 3 (15%) also had rapid shallow sucks during the observation. One (4%) of these infants was ultimately referred to the healthcare provider and transferred to the newborn baby unit. Three (15%) mothers also took their infants off the breast before the infants released the breasts, potentially indicating that the infant was not yet full **(Figure 6)**.

**Figure 6:**
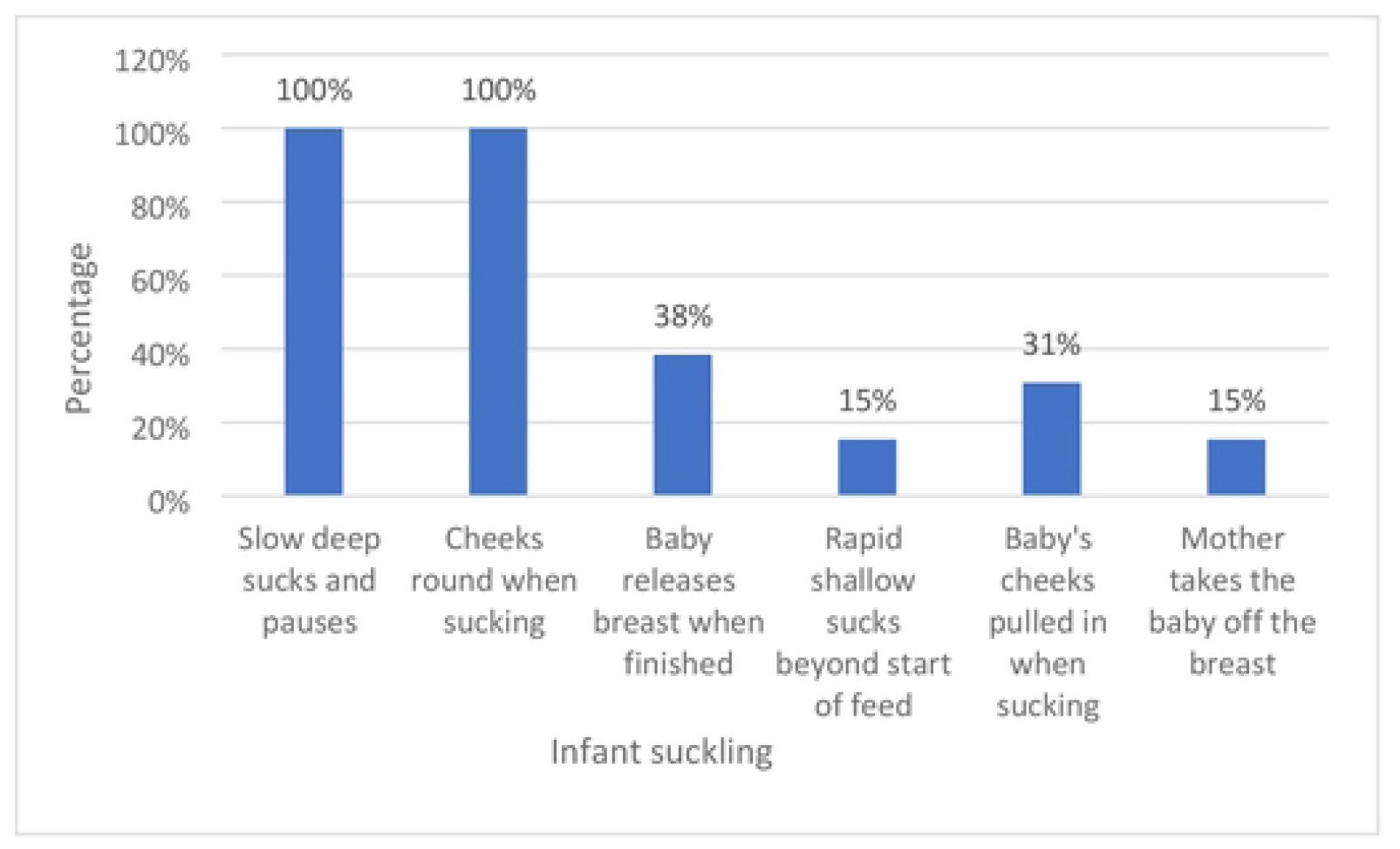
Infant suckling on the breast

### Qualitative findings

Quotes in italics below are from the transcripts and are labelled ‘M’ for LBW mothers, ‘P’ for peer-mothers, and ‘HCW’ for healthcare providers. They are followed by the number of the interview and whether the interview was conducted pre- or post-intervention.

#### Mothers of LBW infants

Among the mothers of LBW infants, key themes were **the challenges of LBW infant feeding decisions**, where often their preference for breastfeeding was hindered by concerns about the inadequacy of breast milk for their infant’s growth and well-being.

Most of the mothers of LBW infants chose to breastfeed their infants, although if their infant was having difficulties with breastfeeding or not gaining weight, mothers reported that healthcare providers would recommend supplementing breast milk with artificial formula milk. Nevertheless, the women often found breastfeeding to be the most convenient method of feeding and had the additional benefit of not being a financial burden on the family.

> *“I was told (by healthcare providers) that being that they were not able to breastfeed after they were born, then I should give them Nan [artificial formula milk] to sustain them” (M11, post-intervention)*
>
> *“Sometimes you don’t have money but being that the mother wants the best for her child she will get motivated [to breastfeed]” (H3, pre-intervention)*

The stigma associated with having a LBW infant in the community also influenced mothers’ infant feeding decision. Under pressure from family members and the community for their infants to gain weight as quickly as possible, family members gave them supplementary feeds, including artificial formula milk, cow’s milk or dilute maize meal porridge.

> *“They just decide that my baby is not getting enough, and so they start to give other feeds like milk from a cow or porridge” (M13, post-intervention)*

The **challenges of feeding LBW infants** included physical limitations, particularly the infant becoming too tired to feed or choking during feeds. These caused fear and anxiety among the mothers, particularly that the baby might choke and die. Expressing breastmilk when their infants were not able to breastfeed was a challenge for mothers of LBW infants. The key problems they reported were pain and difficulties in (manual) expressing, difficulties with storage as many of them did not have access to refrigerators either in the hospital or at home, and the additional time needed. Cultural and religious beliefs also hindered women from expressing breastmilk because this practice was associated with women who had experienced a stillbirth or neonatal death, to relieve breast engorgement. Mothers, therefore, feared that by practising this, they might be blamed for harming their LBW infants

> *“Some people express breastmilk, and the container they use is not that clean, or maybe she left it there and got contaminated” (M14, post-intervention)*
>
> *“I would not do that [express] because it is not recommended in our culture for a living baby” (M12, post-intervention)*

All of the mothers appreciated the advice and support they received from the peer-mothers. Some were advised by the nursing staff on how to breastfeed a LBW infant, but the peer-mothers had more time to sit with the LBW mothers whilst they were feeding, giving more practical support.

> *“I am thankful for teaching me how to breastfeed, I can only thank you for taking your time to teach me” (M11, post-intervention)*
>
> *“A nurse taught me but the information was not rich as the one I got from the peer-mother*.*” (M14, post-intervention)*

The **support from the peer-mothers also gave the women confidence that they could breastfeed their LBW infants**, that there was additional support available if needed, and for some, the knowledge to resist the well-intentioned but ill-informed advice from others in the community, particularly around supplemental feeding and expressing breastmilk. The advice and support were also seen by the LBW mothers, to have positive outcomes for their infants, such as one mother, who said:

> *“She gave me advice and that is why I have improved and if it were not for them, I would not have improved this much, they supported me well” (M11, post-intervention)*

#### Peer-mothers

Peer-mothers across the eight facilities generally felt that the **healthcare providers valued their support and they, therefore, worked collaboratively as a team**. Healthcare providers helped the peer-mothers to identify mothers of LBW infants and were supportive. However, there were instances where the peer-mothers felt that the healthcare providers were agitated by their requests for support and attributed this to the pressure they work under due to being short-staffed.

> *“They helped us identify the LBW babies, they helped us in communication and welcomed us” (P12, post-intervention)*
>
> *“You would find some [HCWs] with bad moods, they could reply rudely but we did understand them” (P13, post-intervention)*

The **combination of the breastfeeding support training and their previous experience of caring for a LBW infant gave the peer-mothers a good basis and confidence to support the LBW mothers**. They were aware however that this was not only advantageous for the LBW mothers in the study but also members of the wider community.

> *“I was taught how to breastfeed the baby, … and with the experience I had of a low birth weight (baby), whenever I went to teach a mother with a LBW baby, I knew what I was doing” (P13, post-intervention)*
>
> *“It helped me a lot and I see that it is also helping the community too because when I teach a mother, she can also get a chance of teaching that mother who did not get the teachings, and you find that the entire community is made aware” (P11, post-intervention)*

The peer-mothers appreciated being able to provide practical advice, such as practising skin-to-skin and KMC, but these efforts were hampered by the difficulties of working with the LBW mothers whilst minimising any risk from Covid-19.

> *“Kangaroo mother care [was difficult] as some mothers did not have leso (a cloth worn by women) to demonstrate the same and because of covid they could not share so I had to use other clothes*.*” (P12, post-intervention)*

#### Healthcare providers

The **healthcare providers generally appreciated the help from the peer-mothers**. As they were understaffed, they acknowledged that they could not spend as much time as necessary with new mothers, who might be having difficulties feeding their newborns. This was particularly the case for those with LBW infants, a gap which the trained peer-mothers were able to, at least partly, fill.

> *“This is the biggest challenge, we are very few and even hence we are not able to help low birth weight infants as we should” (HCW 1, pre-intervention)*
>
> *“While she [peer-mother] is sat there, it becomes helpful because when the mother is not doing the right thing the peer-mother will correct, so it is easy for the mother to remember” (HCW 11, post-intervention)*

## Discussion

This study found that the provision of peer-mother breastfeeding support to mothers of LBW infants was feasible and generally well accepted by both mothers and healthcare providers. They were able to provide advice, encouragement and practical support to the new mothers in hospital, on various aspects of breastfeeding in relation to low birth weight infants. These included positioning and attachment at the breast, hygiene, expressing breast milk and cup feeding.

The benefits of using peer-mothers to support hospital-based breastfeeding interventions have been previously documented. A systematic review by Olufunlayo et al [30]. showed that larger effects were obtained in LMICs from interventions of peer-mothers or lay people providing breastfeeding support in a hospital setting, in combination with existing healthcare providers [30]. However, the benefits of peer-support breastfeeding interventions are less clear following discharge back into the community. In a recently completed study, which sought to explore a community-based package of interventions for LBW infants post-discharge from hospital care in western Kenya, using trained peer-mothers, breastfeeding difficulties and associated sub-optimal infant feeding practices were the greatest challenge that peer-mothers encountered [16]. This was largely due to early discharge of LBW infants before breastfeeding was fully established because of overcrowding and staff shortages on the postnatal wards [16].

Other studies which explored the use of peer-mothers to support mothers of severely malnourished infants under 6 months in healthcare facilities [17] and in the community [31], found that the intervention was a successful strategy for re-lactation and re-establishment of exclusive breastfeeding in the short term, but was often not sustained following discharge from hospital. The authors of the Fadnes et al study hypothesised that reasons for the lack of benefit from their breastfeeding support intervention might have been the inappropriate use of exclusive breastfeeding (for LBW infants who had difficulties breastfeeding, or for infants > 6 months old) or that peer-counsellors, stressing the advantages of breastfeeding might have given the women a false sense of security, thus reducing appropriate health seeking behaviour for common infectious diseases, including malaria and pneumonia [31].

The use of peer-mothers to provide hospital-based breastfeeding support, as in our study, can be beneficial and could maximise the opportunities for teaching and promotion of infant feeding practices in the short time that women are in hospital, particularly for the more vulnerable LBW infants. However, further research is needed to determine the extent to which breastfeeding is continued following discharge from hospital by a combined hospital and community-based peer-mother support approach. Lactation support is a core aspect of postnatal care and in the context of staff shortages, opportunities to embed peer support into the healthcare system could enhance optimal breastfeeding practices and provide continuity of care (hospital-community).

During the interviews, mothers of LBW infants expressed the challenges that they faced in implementing the recommended infant feeding practices in the community, particularly in relation to expressing breastmilk and exclusive breastfeeding. In particular, expressing breastmilk was taboo and associated with relieving breast engorgement after the death of an infant. There was also stigma related to having a LBW infant, with pressure from family members to give the infant non-breastmilk or formula alternatives, such as maize porridge. These findings were also reported by Van Ryneveld et al [32] in Kenya, who suggested the need for less medicalised (or prescriptive) breastfeeding support and the promotion of breastfeeding practices that can be sustained in the community, particularly targeting other caregivers (such as grandparents).

As part of the breastfeeding support intervention, the peer-mothers were taught about the benefits of KMC. This intervention has been found to significantly reduce neonatal mortality among preterm, very LBW infants in hospitals in South Asia and sub-Saharan Africa by 25% [33]. Similarly, in India, community-initiated KMC reduced neonatal mortality among LBW infants by about 30% and improved exclusive breastfeeding rates at 28 days and 3 months [34]. In our study, there were mixed responses about its use, with some finding it helpful whilst others found it a challenge. As our intervention was hospital-based and conducted during the COVID-19 pandemic, infection control measures restricted its full implementation, but it was felt that this warrants further exploration.

The peer-mothers involved in this study underwent a thorough training course to equip them with the knowledge and skills they needed to provide the breastfeeding support interventions, and the results showed that they were very good at assessing mother-infant’s wellbeing. However, they appeared to be less confident with the more technical aspects of breastfeeding, such as observing infant attachment and suckling. As the peer-mothers were only required to have completed basic education and had no prior training in breastfeeding other than to have breastfed their own LBW infant, the provision of additional mentoring on the practical aspects of breastfeeding is required to build and sustain their confidence in assessing this.

### Study limitations

This study has provided valuable insights into the feasibility and acceptability of using peer-mothers to provide breastfeeding support to mothers of LBW infants in hospitals in rural western Kenya. It demonstrated that the intervention could be beneficial and was acceptable to mothers of LBW infants as well as supporting the work of existing healthcare providers. However, because this study was conducted during the COVID-19 pandemic, mothers often had limited support from other family members in the hospitals due to limited or no-visiting policies, and peer-mothers could not access other family members to share knowledge and advice relating to breastfeeding. It also meant that women were discharged earlier from the healthcare facilities, limiting the time during which they had contact with peer-mothers and healthcare providers. Although personal protective equipment was available, peer-mothers additionally had to make difficult decisions regarding their own health, balancing the risk time spent in high-risk hospital settings, with supporting the LBW mothers.

Another challenge faced during the study was the healthcare providers’ strike across Homa Bay County, during which time maternity and neonatal units were closed [35]. This resulted in a four-week delay in the implementation of the intervention, with implications for the retention of knowledge and skills by peer-mothers, study budget, engagement with the healthcare facilities, and recruitment of new mothers.

## Conclusion

Our study found that using trained peer-mothers to provide facility-based breastfeeding support for mothers of LBW newborn infants was both feasible and acceptable, even in the context of the COVID-19 pandemic. However, ongoing support and mentorship for the peer-mothers are required for them to build sustained confidence in providing breastfeeding support. The study demonstrated that deploying peer-mothers, following appropriate training, to provide breastfeeding support to mothers of LBW infants, has the potential to improve the uptake and continuity of appropriate infant feeding practices in rural Western Kenya. This in turn can result in better physical and psychological health outcomes for vulnerable LBW infants, their mothers and families, as well as changing attitudes and reducing stigma in the wider community. These data will inform the design of a larger study that seeks to rigorously evaluate the use of peer-mothers in improving LBW infant feeding and care practises in the postnatal period, both in hospital and post-discharge, in rural communities in sub-Saharan Africa.

## Data Availability

Data used in this study is provided as part of the submitted article.

## Acknowledgements

The study authors would like to gratefully acknowledge the help and contribution of the six peer-mothers who helped with this study: Beatrice Auma John, Hilda Awino Owidi, Linda Auma Ogondi, Malath Akinyi Nyagori, Maureen Achieng Ouma & Merlin Beshna Oyudo.

The authors of this study also appreciate the support given by the staff from each of the included facilities, especially: Beatrice Akinyi, Homa Bay; Mervine Agung, St. Paul Hospital; Lucas Okoyo, Nyalkinyi Hospital; Leonard Walela, Nyamasi Hospital; Monica Obel, St. Lawrence Hospital; Nick Were, Pala Hospital; Tobias Ochieng, Marindi Hospital; Timothy Onyango, Samarimed Hospital.

## References

1. UN-IGME. Levels and trends in child mortality report. 2021; Available at: https://www.who.int/publications/m/item/levels-and-trends-in-child-mortality-report-2021

2. United Nations (2022) The Sustainable development Goals Report. 2022; Available at: https://sdgs.un.org/goals#implementation

3. UNICEF-WHO. Low birthweight estimates: Levels and trends 2000-2015. 2019; Available at: https://www.who.int/nutrition/publications/UNICEF-WHO-lowbirthweight-estimates-2019/en/

4. Lawn JE, Cousens S, Zupan J, The Lancet Neonatal Survival Steering Team. 4 million neonatal deaths: when? Where? Why? Lancet. 2005; 365(9462):891–900.

5. Aluvaala J, Okello D, Murithi G, Wafula L, Wanjala L, Isika N, et al. Delivery outcomes and patterns of morbidity and mortality for neonatal admissions in five Kenyan hospitals. J Trop Pediatr. 2015; 61(4):255–259.

6. O’Leary M, Edmond K, Floyd S, Newton S, Thomas G, Thomas SL. A cohort study of low birth weight and health outcomes in the first year of life, Ghana. Bull World Health Organ. 2017; 95(8):574–583.

7. Gladstone M, Oliver C, Van den Broek N. Survival, morbidity, growth and developmental delay for babies born preterm in low and middle income countries - a systematic review of outcomes measured. PLoS One. 2015; 10(3):e0120566.

8. WHO, UNICEF. Survive and Thrive: Transforming the care of every small and sick newborn. Geneva: WHO; 2019.

9. UNICEF, WHO. Countdown to 2030: tracking progress towards universal coverage for reproductive, maternal, newborn, and child health. Lancet. 2018; 391(10129):1538–1548.

10. World Health Organization. WHA Global Nutrition Targets 2025: Low Birth Weight Policy Brief. Geneva: WHO; 2014.

11. Bhutta ZA, Das JK, Bahl R, Lawn JE, Salam RA, Paul VK et al. Can available interventions end preventable deaths in mothers, newborn babies, and stillbirths, and at what cost? Lancet. 2014; 384(9940):347–370.

12. World Health Organization. WHO Recommendations on postnatal care of the mother and newborn. Geneva: WHO; 2013.

13. World Health Organization, UNICEF. Baby-friendly hospital initiative: Revised, Updated and Expanded for Integrated Care. Geneva: WHO; 2009.

14. Upadhyay RP, Martines JC, Taneja S, Mazumder S, Bahl R, Bhandari N, et al. Risk of postneonatal mortality, hospitalisation and suboptimal breast feeding practices in low birthweight infants from rural Haryana, India: findings from a secondary data analysis. BMJ Open 2018; 8(6):e020384.

15. Rotich E, Wolvaardt L. A descriptive study of the health information needs of Kenyan women in the first 6 weeks postpartum. BMC Pregnancy Childbirth 2017; 17(1):385.

16. Were F, Barsosio H, Juma D, Ayaye E, Omondi E, Boga M, et al. The feasibility of using peer mothers to deliver a community-based package of interventions to low birth weight infants post discharge from hospital care in Homa Bay County, Kenya. In: 10th KEMRI Annual Scientific and Health conference: 2020. KEMRI; 2020: 395.

17. Mwangome M, Murunga S, Kahindi J, Gwiyo P, Mwasho G, Talbert A, et al. Individualized breastfeeding support for acutely ill, malnourished infants under 6 months old. Matern Child Nutr. 2020; 16(1):e12868.

18. Kenya National Bureau of Statistics, Ministry of Health, National AIDS Control Council, Kenya Medical Research Institute, National Council for Population and Development, International, ICF International. Kenya Demographic and Health Survey 2014. Available at: https://dhsprogram.com/publications/publication-fr308-dhs-final-reports.cfm

19. National AIDS and STI Control Programme (NASCOP). Kenya Population-based HIV Impact Assessment (KENPHIA) 2018: Final Report. Nairobi: NASCOP; 2022. Available at: https://phia.icap.columbia.edu/kenya-final-report-2018/

20. World bank. Mortality rate, neonatal (per 1,000 live births). 2020. Available at: https://data.worldbank.org/indicator/SH.DYN.NMRT

21. Kenya National Bureau of Statistics. Nyanza Province Multiple Indicator Cluster Survey 2011, Final Report. Nairobi: Kenya National Bureau of Statistics; 2013.

22. Creswell JW: Research design: qualitative, quantitative, and mixed methods approaches. New York: Sage; 2014.

23. Ministry of Public Health and Sanitation, Kenya. National Strategy on Infant and Young Child Feeding. 2007. Available at: https://extranet.who.int/nutrition/gina/en/node/8537

24. World Health Organisation. WHO Guide: Counselling for women to improve breastfeeding practices. Geneva: WHO; 2018.

25. The Global Health Network. Guide to the iCARE-Haaland Model Resources. Available at: https://connect.tghn.org/training/icare-haaland-model/guide-icare-haaland-model-resources/

26. Otiti MI, Kariuki S, Wang D, Hall LJ, Ter Kuile FO, Allen S. PRObiotics and SYNbiotics to improve gut health and growth in infants in western Kenya (PROSYNK Trial): study protocol for a 4-arm, open-label, randomised, controlled trial. Trials. 2022; 23(1):284. doi: 10.1186/s13063-022-06211-1

27. Marshall MN. Sampling for qualitative research. Fam Pract 1996, 13(6):522–525.

28. Fusch PI, Ness LR. Are We There Yet? Data Saturation in Qualitative Research. Qual Rep. 2015; 20(9):1408–1416.

29. Kiger ME, Varpio L. (2020) Thematic analysis of qualitative data: AMEE guide no. 131. Med Teach. doi: 10.1080/0142159X.2020.1755030

30. Olufunlayo TF, Roberts AA, MacArthur C, Thomas N, Odeyemi KA, Price M, et al. Improving exclusive breastfeeding in low and middle-income countries: A systematic review. Matern Child Nutr. 2019; 15(3):e12788.

31. Fadnes LT, Nankabirwa V, Engebrestsen IM, Sommerfelt H, Birungi N, Lombard C, et al. Effects of an exclusive breastfeeding intervention for six months on growth patterns of 4-5 year old children in Uganda: the cluster-randomised PROMISE EBF trial. BMC Public Health. 2016; 16(555)

32. Van Ryneveld M, Mwangome M, Kahindi J, Jones C. Mothers’ experiences of exclusive breastfeeding in a postdischarge home setting. Matern Child Nutr. 2020; 16(4)

33. Arya S, Naburi H, Kawaza K, Newton S, Anyabolu CH, Bergman N, Rao SPN, Mittal P, Assenga E, Gadama L et al: Immediate “Kangaroo Mother Care” and Survival of Infants with Low Birth Weight. N Engl J Med 2021, 384(21):2028–2038.

34. Mazumder S, Taneja S, Dube B, Bhatia K, Ghosh R, Shekhar M, et al. Effect of community-initiated kangaroo mother care on survival of infants with low birthweight: a randomised controlled trial. Lancet. 2019; 394:1724–36

35. Mohiddin A, Langat E, Orwa J, Naanyu V, Temmerman M. Exploring the impact of health worker strikes on maternal and child health in Kenyan county. BMC Health Serv Res. 2022; 22(1)

